# Clinical validity of expanded carrier screening: evaluating the gene-disease relationship in more than 200 conditions

**DOI:** 10.1101/2019.12.13.19014894

**Authors:** Marie Balzotti, Linyan Meng, Dale Muzzey, Katherine Johansen Taber, Kyle Beauchamp, Myriad Genetics Curation Team, Baylor Genetics Curation Team, Rebecca Mar-Heyming, Bethany Buckley, Krista Moyer

**Affiliations:** Myriad Genetics, South San Francisco, CA; Baylor Genetics, Houston, TX; Tempus, Redwood City, CA; Ambry Genetics, Aliso Viejo, CA; Invitae, San Francisco, CA

## Abstract

**Objective:** Clinical guidelines consider expanded carrier screening (ECS) to be an acceptable method of carrier screening. However, broader guideline support and payer adoption require evidence for associations between the genes on ECS panels and the conditions for which they aim to identify carriers. We applied a standardized framework for evaluation of gene-disease association to assess the clinical validity of conditions screened by ECS panels.

**Methods:** The ClinGen gene curation framework was used to assess genetic and experimental evidence of associations between 208 genes and conditions screened on two commercial ECS panels. Twenty-one conditions were previously classified by ClinGen, and the remaining 187 were evaluated by curation teams at two laboratories. To ensure consistent application of the framework across the laboratories, concordance was evaluated on a subset of conditions.

**Results:** All 208 evaluated conditions met the evidence threshold for supporting a gene-disease association. Furthermore, 203 of 208 (98%) achieved the strongest (“Definitive”) level of gene-disease association. All conditions evaluated by both commercial laboratories were similarly classified.

**Conclusion:** Assessment using the ClinGen standardized framework revealed strong evidence of gene-disease association for conditions on two ECS panels. This result establishes the disease-level clinical validity of the panels considered herein.

## Introduction

A genetic test has clinical validity if it accurately detects genetic variations associated with clinical outcomes of interest. For screening panels comprised of tens to hundreds of single-gene disorders, clinical validity is demonstrated, in part, by evidence of association between each gene and its ostensibly associated condition. Clinicians offering testing to their patients must have confidence that the role of a gene in disease causation is well established so that accurate diagnoses can be made and appropriate management undertaken based on the test result.

Evidence demonstrating clinical validity is often a foundational requirement for clinical guidelines to support genetic testing (Grody et al., 2013;; National Academies of Sciences Engineering and Medicine, 2017). Similarly, insurers cite the lack of clinical validity evidence as a reason for non-coverage of some genetic tests (Chambers et al., 2017).

The purpose of carrier screening is to determine whether couples are at high risk of having children affected with serious genetic conditions, allowing for informed reproductive decision-making and pregnancy management. The number of gene-disease pairs currently available for carrier screening varies across laboratories, with some offering screening for only a few conditions and others offering screening for hundreds (termed expanded carrier screening or ECS). Many factors influence the selection of conditions that are included on ECS panels, such as the severity of the condition and its prevalence in the population, as well as the screen’s clinical sensitivity for pathogenic variants (Beauchamp et al., 2018;; Ben-Shachar et al., 2019;; Kaseniit et al., 2019). Regardless of the differences in the sets of conditions screened by commercial ECS panels, all conditions on a given panel should have established clinical validity at the level of gene-disease association because important reproductive and pregnancy management decisions are made based on results. However, to our knowledge, no systematic evaluations of gene-disease associations on ECS panels have been undertaken.

The Clinical Genome Resource (ClinGen) (Rehm et al., 2015) has developed a framework to classify gene-disease relationships by the quantity and quality of the evidence supporting such a relationship (Strande et al., 2017). The framework provides a system by which supporting and contradictory gene-disease association evidence can be organized and then categorized by strength, resulting in a quantification of clinical validity for the gene-disease relationship being interrogated. Evidence classifications that are supportive of an association include “Definitive,” “Strong,” “Moderate,” and “Limited.” Contradictory classifications include “Disputed” and “Refuted.” A classification of “No reported evidence” indicates no reported pathogenic or likely pathogenic variants in humans.

The ClinGen gene-disease association framework has been applied in a number of disease-specific areas to determine the clinical validity of genes included on panels (DiStefano et al., 2019;; Hosseini et al., 2018;; Ingles et al., 2019;; Lee et al., 2019;; McGlaughon et al., 2019;; Renard et al., 2018;; Seifert et al., 2019). For example, of 21 genes included on panels assessing single-gene causes of Brugada syndrome, an arrhythmia condition that increases risk for sudden cardiac death, only one gene (*SCN5A*) was classified as having Definitive evidence of clinical validity, while the remaining 20 were classified as Disputed (Hosseini et al., 2018). The study concluded that more evidence was needed before genes other than *SCN5A* are tested as the cause of Brugada syndrome (Hosseini et al., 2018). Another study applied the framework to 31 genes associated with hereditary cancer syndromes (Lee et al., 2019). The study classified the evidence as supportive for 25 genes (Definitive for 18, Moderate for 1, and Limited for 6), and as contradictory (Disputed) for three genes;; it found no reported evidence for two genes. The authors suggested that their results could be used to optimize the design of hereditary cancer panels (Lee et al., 2019).

In this study, we have used the ClinGen framework to assess the clinical validity of the gene-disease relationship for more than 200 autosomal recessive and X-linked conditions included on commercially available ECS panels.

## Methods

### Editorial Policies and Ethical Considerations

This study did not involve human participants and therefore was not subject to ethical review or institutional review board approval.

### Gene Curation Framework

The ClinGen gene curation framework provides a method to evaluate the strength of a gene-disease association (Strande et al., 2017). According to this framework, papers published in peer-reviewed journals are curated for different evidence types, specifically genetic and experimental evidence. Genetic evidence includes case-level data (e.g., studies describing individuals with variants in the gene of interest and segregation analysis) and case-control data. Experimental evidence is grouped into three main categories: functional evidence (e.g., biochemical, protein interactions, expression), functional alteration evidence (e.g., the gene or gene product function is altered in patient or non-patient cells with candidate mutations), and models/rescue evidence (e.g., replication of the disease in a model organism or model system, and rescue of the disease phenotype with wild-type gene or gene product). Curated genetic and experimental evidence from the literature is gathered and assigned points;; higher point values reflect stronger support for the association (see Gene Curation below). For example, model/rescue experiments are assigned higher default points (1-2 points) than function experiments (0.5 points). The maximum number of points obtainable for genetic and experimental evidence is 12 and 6 points, respectively. Following data collection, annotation and summing the evidence points, one of seven possible clinical validity classifications is assigned to each gene-disease pair. Supportive classifications are based on the total point sum and qualify the level of support for the association: “Definitive” (12-18 points plus replication over time, i.e., three or more independent publications with convincing evidence over a time frame greater than three years), “Strong” (12-18 points without replication over time), “Moderate” (7-11 points), and “Limited” (1-6 points). Further, a classification of “No Reported Evidence” is applied to gene-disease pairs for which there are no reports of patients with the disease having pathogenic or likely pathogenic variants in the gene. Two classifications exist within the framework for gene-disease associations with contradictory evidence: “Disputed” is applied when valid contradictory evidence has arisen since the initial gene-disease association claim, and “Refuted” is applied when the gene-disease association has been fully refuted by valid contradictory evidence since the time of the original claim.

### Gene Curation

The ClinGen framework was applied to 208 gene-disease pairs included in commercially available ECS panels by Myriad Women’s Health (Foresight) and Baylor Genetics (GeneAware). The 208 gene-disease pairs were curated independently at Myriad and/or Baylor as described below and as listed in Supplementary Table 1. Each gene curation team consisted of gene curators and a laboratory director. Gene curators (who are specially trained scientists with Masters and/or PhD degrees) followed the ClinGen Gene Curation Standard Operation Procedure, Version 5 (The Clinical Genome Resource Gene Curation Working Group, 2017). Curation could be discontinued once a “Definitive” classification was reached: i.e., once ≥12 points of genetic evidence had been amassed and at least two pieces of experimental evidence had been curated from two or more independent papers (non-overlapping clinical studies produced by different groups). Each gene-disease pair was curated by a single curator, and then laboratory directors reviewed all gene curations for appropriate application of the framework and point assignments;; evaluated contradictory evidence, if any;; assessed consistency;; recommended changes when needed;; and approved gene curations that met review criteria. A set of 21 gene-disease pairs were curated independently by both Myriad and Baylor to determine the level of classification concordance between the two institutions. These gene-disease pairs were chosen to represent both average difficulty curations (those with clear case and experimental data or clear mechanism of action, for example) and challenging curations (those with few cases or experimental studies, for example). Gene-disease pairs with conflicting classifications between the two institutions were examined, evidence was combined where applicable, and the appropriate assignment of points was discussed between Myriad and Baylor laboratory directors until a concordant classification was reached. ClinGen gene curation expert panels had previously curated and assigned clinical validity classifications to an additional 21 gene-disease pairs that were included in the ECS panels evaluated in this study (DiStefano et al., 2019;; McGlaughon et al., 2019;; The Clinical Genome Resource);; we did not reapply the framework to these 21 genes (Supplementary Table 1). In addition to the 208 conditions included on Myriad’s and Baylor’s ECS panels, Myriad also evaluated nine recessive conditions not typically screened during expanded carrier screening (Supplementary Table 2). All classifications have been submitted to ClinGen for public availability.

**Table 1.** Gene-disease association strengths for the 208 routine ECS conditions and the nine that approximated “negative controls.”

|  | Definitive | Strong | Moderate | Limited | No Evidence | Disputed | Refuted | Total |
| --- | --- | --- | --- | --- | --- | --- | --- | --- |
| **Carrier Screen Panel Gene Set** | 203 | 0 | 4 | 1 | 0 | 0 | 0 | 208 |
| **“Negative Control” Gene Set** | 1 | 2 | 4 | 2 | 0 | 0 | 0 | 9 |

## Results

The gene-disease relationship of each of 208 conditions included on two commercially available ECS panels was evaluated using the ClinGen framework. In total, more than 2,000 pieces of evidence were included in the final classifications, with each gene-disease pair having a median of 11 pieces of evidence (Figure 1). Of the 208 conditions, 203 (98%) were found to have a Definitive association, the strongest level of evidence for clinical validity (Figure 1, Table 1, Supplementary Table 1). Of the remaining five conditions, four were classified as having Moderate evidence and one was classified as having Limited evidence (Figure 1, Table 1, Supplementary Table 1). Notably, all evaluated conditions were classified into categories in the supportive clinical validity range, i.e., no conditions had Refuted, Disputed, or No reported evidence. To approximate a negative control, we also evaluated nine recessive conditions not routinely screened in a general population, but sometimes screened in select higher-risk groups, such as Ashkenazi Jewish. In contrast to the commonly screened conditions, the nine rare conditions predominantly showed either Moderate or Limited evidence (Table 1, Supplementary Table 2);; these categories are still considered supportive of clinical validity.

**Figure 1.**
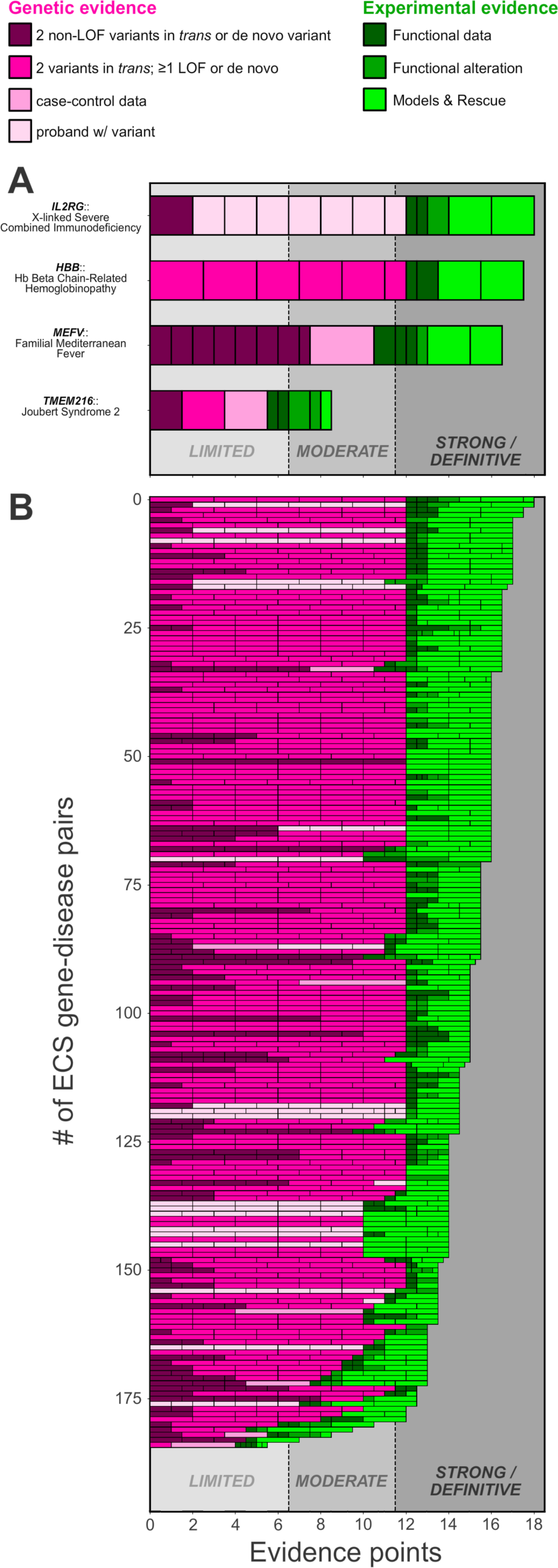
Application of ClinGen framework to expanded carrier screening conditions shows predominantly definitive gene-disease associations. (A) The evidence underlying four representative gene-disease pairs are shown, where each segment of the bar graph indicates the type of evidence (color, see key) and score (width of the segment). Classifications are indicated by the gray-shaded regions. (B) Bar graphs as in (A) are shown for the 193 gene-disease pairs classified by Myriad and/or Baylor. (For the 21 gene-disease pairs classified by both, only the Myriad data is shown for visual clarity.)

Next, we examined the distribution of quantitative genetic and experimental scores (Figure 1, panel A shows close-up examples of the 208 shown in panel B). Along with the replication over time, these scores were used to determine the ClinGen classification of each gene-disease relationship. Most gene-disease pairs had the maximum possible genetic score of 12 (Figure 1A, pink). Experimental scores were more uniformly distributed across the possible range (i.e., the green-shaded bars in Figure 1B range from 0.5 to 6);; however, this may be partially due to downward censorship bias in the experimental data, which occurs because upon reaching the highest classification status (Definitive), a curator may discontinue the experimental literature search. Finally, as a check of concordance across curating laboratories, we compared total scores for the 21 conditions that were jointly curated. Nineteen of the 21 conditions were classified concordantly as Definitive. The other two gene-disease pairs (*SLC35A3* - autism spectrum disorder/epilepsy/arthrogryposis and *TMEM216* - Joubert syndrome 2/Meckel syndrome type 2) were classified discordantly, with Myriad classifying both as Moderate and Baylor classifying both as Definitive (notably, both were in the Supportive evidence category).

After discussion between the two teams, consensus was reached on Moderate classifications for both (Supplementary Table 1).

## Discussion

An ECS has clinical validity if it correctly discovers pathogenic variants in the panel’s genes that determine carrier status for the screened conditions. Accordingly, evidence of the following is required to demonstrate clinical validity: (1) the ECS accurately identifies variants in genes of interest, (2) the ECS correctly classifies the discovered variants as pathogenic or not, and (3) pathogenic variants in the genes of interest are associated with carrier status of the panel’s conditions. The first has been demonstrated via analytical validation of a large ECS panel (Hogan et al., 2018), where sensitivity and specificity were both >99% for SNVs and indels. The second was demonstrated by the comparison of variant classification concordance between a commercial ECS offering and the consensus in ClinVar (Kaseniit et al., 2019);; notably, classifications were 99% concordant, with fewer than 1 in 500 of those tested expected to carry a variant with disputed classification. The third is addressed in this study, which used a widely accepted and standardized framework to show that >97.5% of genes screened by the superset of two large ECS panels are definitively associated with the specified condition. Taken together, these three lines of evidence strongly establish the clinical validity of ECS.

Several important considerations are prompted by our study. First, many criteria should be assessed when implementing ECS, and having clinical validity is not sufficient reason alone to include a condition on an ECS panel (Henneman et al., 2016). For instance, the severity, clinical utility, and prevalence of screened conditions are all important factors (Beauchamp et al., 2018): mild or exceedingly rare conditions may not warrant inclusion, even if gene-disease association is clear. Second, although the 208 genes studied here were almost entirely classified as having Definitive clinical validity evidence, such supportive classifications may not generalize to extremely large panels;; thus, it is incumbent upon ECS laboratories to undertake clinical validity evaluations when designing panels. Third, we anticipate that larger panels that screen for very rare conditions may have unestablished clinical validity; eight of nine (89%) rare conditions analyzed as “negative controls” in this study had less than Definitive gene-disease association, as compared to only 2.5% with non-Definitive evidence for conditions on the panels.

A number of different multi-gene panel tests, including those assessing hereditary cancer syndrome risk, cardiac and thoracic condition diagnosis, and underlying causes of hearing loss, have been evaluated by application of the ClinGen clinical validity framework (DiStefano et al., 2019;; Ingles et al., 2019;; Lee et al., 2019;; Renard et al., 2018;; Seifert et al., 2019). The ability of the ClinGen framework to be applied in an evidence-based and systematic manner in many different disease areas is a testament to its strength as a tool for clinical validity assessment of monogenic conditions. Additionally, its decisive scoring mechanism controls for potential subjectivity among curators as to the weight of different types of evidence. Indeed, in this study, 19/21 (90%) of classifications were concordant, and all showed supportive evidence for a gene-disease association.

As new evidence emerges, gene-disease association classifications may change, particularly for those with lower point totals at initial curation, such as Limited, No Reported Evidence, or Contradictory Evidence categories. In this study, one gene, *HYLS1* (associated with hydrolethalus syndrome), was classified as having Limited evidence, the lowest of the supportive categories. In several studies applying the ClinGen framework, discussion of genes classified as Limited have noted the need to regularly revisit curations so that new evidence can be assessed (Grant et al., 2018;; McGlaughon et al., 2019;; Renard et al., 2018). McGlaughon et al. (2018) retrospectively analyzed classifications of 22 gene-disease pairs curated yearly, and found that gene-disease associations at the low end of the Limited point range (< 2 points) are more likely to remain Limited or become Disputed/Refuted than than those initially scored in mid to upper Limited point range (2–6 points) (McGlaughon et al., 2018). Notably, *HYLS1* was scored at 5.5 points, suggesting that it will remain in the supportive clinical validity category. Nonetheless, it, like other genes being screened by ECS, should be subject to reassessment as new evidence emerges.

Limitations to this study should be noted. First, though the ClinGen framework should result in the same classification even when applied by different curators, we cannot guarantee that other curators would have categorized the conditions identically to this study. Second, and related to the first, one curation team independently categorized more conditions than the other;; if a bias toward stronger or weaker classifications existed in the team that completed more curations, it may have skewed the results. However, we checked the consistency of curations between the two teams by curating an overlapping set of 21 conditions, and found >90% initial concordance (and eventual consensus on the remainder). And third, the categorizations represent the state of the evidence at the time that the curations were completed. As noted previously, additional evidence may emerge over time, changing the classification;; however, due to the already rigorous evidence requirements for supportive classifications, additional evidence is likely to increase the strength of the association, rather than weaken it (McGlaughon et al., 2018).

Our study showed that the vast majority of conditions on the ECS panels explored herein have definitive support for gene-disease association, a critical component of ECS clinical validity. Established clinical validity is necessary to provide clinicians with the confidence that test results are accurate indicators of disease causation. This is especially important in the carrier screening setting, in which results guide reproductive and pregnancy management decisions.

## Data Availability

All gene-disease associations have been submitted to ClinGen for public posting.

## Acknowledgements

The authors thank Jim Goldberg for critical review of the manuscript and Anna Gardiner for assistance in manuscript preparation.

## Consortia

### Myriad Genetics Curation Team

David Tran, Jessica Conner, Megan Li, Martha Arnaud, Lynne Nazareth, Kerri Hensley, Michael Hunter, Sam Cox, Domenic Previte, Brianna Campbell, Katrina Tanaka, Candy Heyen, Kaitlin Sesock, Srikanth Appikonda, Ildiko Thibodeau, Majesta O’Bleness, Matthew Brown, Caiqian Cropper, Brandee Price, Juliette Kahle, Kamalika Moulick, Jessica Ray, John Castiblanco, Lea Gemmel, Lisa Spock, Megan Judkins

### Baylor Genetics Curation Team

Blake Atwood, Shen Gu, Ye Cao, Rajarshi Ghosh, Bo Yuan, Fan Xia

## Data Availability Statement

All curations have been submitted to ClinGen for public availability. All data are additionally available from the corresponding author upon request.

